# Temporal Patterns and Association Between FIFA World Cup Tournament Periods and Emergency Department–Treated Injuries

**DOI:** 10.64898/2026.07.10.26357721

**Authors:** Renduchinthala Sai Praneeth Kumar, Jiancheng Ye

## Abstract

**Background:** Major soccer tournaments may temporarily change recreational soccer activity, community gatherings, and injury-prevention needs, but evidence for population-level emergency department (ED) injury patterns during these events is limited. Understanding whether ED-treated soccer injury burden changes during Men’s FIFA World Cup periods may help inform surveillance readiness and prevention planning for future tournaments.

**Objective:** To evaluate whether Men’s FIFA World Cup tournament periods temporally coincided with changes in ED-treated soccer-coded injury burden in the United States and to assess the implications for public health surveillance and injury-prevention preparedness.

**Methods:** We conducted a retrospective, repeated cross-sectional calendar-period analysis of publicly available national ED injury surveillance records from 1999 through 2025. Soccer-coded injuries were identified using product code 1267 in any available product field. The primary exposure was the set of official Men’s FIFA World Cup tournament dates from 2002, 2006, 2010, 2014, 2018, and 2022. Tournament dates were compared with matched same-calendar dates in adjacent years, excluding dates that overlapped other FIFA World Cup tournament windows. The primary estimands were the mean daily difference and ratio in weighted national ED-treated soccer-coded injury estimates between tournament and matched-control periods.

**Results:** The analytic cohort included 170,679 soccer-coded ED cases, corresponding to an estimated 5,366,681 ED-treated soccer-coded injuries nationally. Mean daily weighted estimates were 453.1 during Men’s World Cup tournament dates and 384.1 during matched control dates. The absolute mean daily difference was 68.9 injuries per day (95% CI, -0.5 to 138.3), and the mean daily ratio was 1.18 (95% CI, 1.00 to 1.39). Tournament-specific estimates were heterogeneous, with a near-null estimate for the 2022 winter tournament and higher estimates for prior summer tournaments.

**Conclusions:** Mean daily injury estimates were modestly higher during tournament periods, although the confidence interval for the absolute difference included zero and estimates varied across tournaments. The findings suggest that major soccer events offer valuable opportunities for real-time ED injury surveillance, targeted recreational soccer injury-prevention messaging, concussion awareness, and coordinated public health preparedness for community- and fan-related injury patterns during future tournaments.

## INTRODUCTION

Soccer is among the most widely played team sports in the United States, and soccer-related injuries are an important source of emergency department (ED) utilization, particularly among children, adolescents, and young adults.[1] National surveillance analyses based on the National Electronic Injury Surveillance System (NEISS) have characterized large numbers of ED-treated soccer-related injuries, with sprains and strains, fractures, contusions, and head injuries among the most common diagnoses.[2, 3] Because NEISS captures injuries treated in a probability sample of hospital EDs, such analyses describe ED-treated injury burden represented by the surveillance system rather than total soccer participation injury incidence.[3, 4] More broadly, NEISS has supported national ED surveillance studies across injury and adverse-event topics.[5, 6]

Major international sporting events have been associated with measurable changes in acute health-care utilization in host and viewing populations. Studies conducted around FIFA World Cup tournaments have reported short-term changes in cardiovascular events and hospital admissions during match periods, using event-window designs that compare exposed calendar dates with matched control dates.[7] Standardized injury surveillance within FIFA tournaments has likewise documented injury patterns among elite players.[8, 9] Comparable evidence on ED-treated soccer-coded injuries during men’s World Cup periods in the United States is limited.

Several mechanisms could plausibly contribute to short-term changes in soccer-coded ED injury counts during FIFA World Cup periods. Extensive tournament coverage may increase recreational soccer participation among youth and adults through informal play, organized community events, or pickup matches. Fan gatherings and celebrations may also influence injury patterns. Conversely, increased time spent watching matches could reduce recreational soccer participation and attenuate injury counts. In addition, seasonal variation, school schedules, weather conditions, and local opportunities for soccer participation may differ across tournament periods and contribute to observed temporal patterns.[10]

A tournament-window design is preferable to a calendar-year design for an acute question of this kind, because a World Cup tournament occupies only several weeks of a calendar year; classifying the entire year as exposed dilutes any acute association and increases vulnerability to secular trends.[7, 11] The primary objective of this study was to quantify the ecological calendar-period association between men’s FIFA World Cup tournament dates and NEISS-estimated ED-treated soccer-coded injury counts in the United States from 1999 through 2025, comparing official tournament dates with matched same-calendar adjacent-year control dates; the primary estimands were the mean daily weighted difference and ratio. This surveillance framing is relevant to the 2026 FIFA World Cup and future tournaments,[12] as prior tournament windows can inform monitoring plans and prevention messaging.

## MATERIALS AND METHODS

### Study Design and Data Source

We conducted a retrospective, repeated cross-sectional analysis of publicly available NEISS case-level records from January 1, 1999, through December 31, 2025.[13, 14] NEISS, operated by the United States Consumer Product Safety Commission (CPSC), is a stratified national probability sample of hospital EDs; each sampled case carries a statistical weight, and weighted analyses yield national estimates of ED-treated consumer product and recreation-related injuries.[15] The unit of analysis was the NEISS ED case.

### Source, Database, and Study Populations

The source population comprised persons with consumer product and recreation-related injuries treated in United States hospital EDs and represented by the NEISS sampling frame.[15] The database population comprised all public NEISS records for 1999 through 2025 with valid treatment date, statistical weight, stratum, and primary sampling unit information (9,794,932 records). The study population comprised database records meeting the product-code-defined soccer case definition described below (170,679 records). The denominator was the NEISS-estimated population of ED-treated consumer product– and recreation-related injuries.[15, 16] Of 9,794,932 public NEISS records, all were analysis eligible; 170,679 carried soccer product code 1267, and none were excluded by the plausibility review, yielding a final analytic cohort of 170,679 soccer-coded cases.

### Primary Outcome and Case Definition

The primary outcome was an ED-treated soccer-coded injury identified by NEISS product code 1267 (soccer activity, apparel, or equipment) recorded in any available product field (Product 1 and Product 2 throughout the study period, and Product 3 from 2018 onward, when that field became available).[17] Product-code comparability across the study period was reviewed against CPSC coding documentation before longitudinal analysis, and code 1267 was used as the primary definition.[17] Hereafter, these cases are termed soccer-coded injuries. Because Product 3 was unavailable before 2018, post-2018 ascertainment may have been slightly more complete than earlier ascertainment; this limitation was considered when interpreting post-2018 estimates. Narrative-keyword and combined product-or-narrative definitions were used only as exploratory sensitivity outcomes.[18]

### Exposure and Matched-Control Definitions

The primary exposure was the set of official men’s FIFA World Cup tournament dates within the study period, covering the six tournaments held in 2002, 2006, 2010, 2014, 2018, and 2022 (186 exposed days).[19] Tournament dates were taken from the FIFA event calendar and are reproduced in the supplement.[19] Because NEISS records treatment date rather than injury time or match-viewing time, official tournament dates were mapped to U.S. treatment-date calendar days rather than match kickoff hours. This approach is appropriate for a calendar-period surveillance question but may introduce limited one-day exposure misclassification for tournaments held outside U.S. time zones, especially the 2002 Korea/Japan tournament. The primary comparison comprised matched same-calendar dates in the adjacent year before and the adjacent year after each tournament, excluding dates that overlapped another prespecified men’s or women’s FIFA World Cup tournament period (308 control days). Same-calendar controls were primary because they align month, day, school season, and broad seasonal activity patterns; same-day-of-week controls were analyzed as a robustness check because ED injury volume can vary by weekday. Excluding women’s World Cup windows reduced the possibility of treating another FIFA tournament as an unexposed control period.

### Covariates and Variable Coding

Analysis variables included treatment date, calendar week, year, age, sex, diagnosis, body part, disposition, incident location, product codes, narrative text, statistical weight, stratum, and primary sampling unit. NEISS age coding was harmonized before analysis: ages recorded in months under the NEISS convention for children younger than 2 years (codes 2XX) were converted to years, and age code 0 was classified as unknown.[17] The COVID-19 pandemic period (2020–2021) was retained in the data and addressed with a pandemic-period indicator in adjusted models and with a pandemic-exclusion sensitivity analysis.[20]

### Statistical Analysis

National estimates were calculated using NEISS sample weights. For each descriptive estimate, we reported the unweighted sample count, weighted national estimate, weighted percentage where applicable, design-based 95% CI, and coefficient of variation (CV). Variance estimation used survey-year-specific strata and PSUs in a with-replacement ultimate-cluster approach based on public-use design variables. For tournament-control contrasts, we calculated within-PSU differences between tournament-period and matched-control weighted counts; ratio CIs used log-ratio linearization, preserving within-PSU covariance. PSUs and strata were treated as survey-year-specific identifiers. This public-use paired-contrast implementation extends the CPSC-documented Taylor-series approach to the custom tournament-control contrast, whereas the official CPSC Query Builder primarily provides annual and other standard estimates rather than this paired event-window estimand. Following CPSC screening criteria, estimates were flagged or suppressed when the unweighted count was below 20, the weighted estimate was below 1,200, or the CV exceeded 33%.[16] External calibration against the official CPSC NEISS Query Builder for soccer product code 1267 in 2021–2025 showed that unweighted case counts and weighted national estimates matched CPSC output exactly after rounding (Supplementary Table S7). Confidence-limit endpoints differed slightly because the public-use variance implementation is not identical to CPSC’s internal implementation.

The primary descriptive estimands were the absolute difference and ratio of survey-weighted mean daily national estimates between men’s tournament dates and matched same-calendar controls. Additive and ratio estimates were interpreted together, with emphasis on the range of compatible effects rather than on threshold-based significance. No formal multiplicity adjustment was applied across sensitivity analyses, subgroup analyses, or tournament-specific estimates; these analyses were interpreted as robustness and heterogeneity assessments rather than independent confirmatory tests.

Adjusted analyses were retained as sensitivity analyses because weekly injury series exhibit temporal autocorrelation and conclusions can depend on specification.[7] Weekly national estimates were aggregated to 1,408 complete calendar weeks after excluding 2 incomplete boundary weeks. The main adjusted specification was a survey-variance-weighted AR(1) log-linear WLS model with Newey-West HAC standard errors. The tournament-versus-control ratio was computed as exp(β_tournament_ − β_control_), where β_tournament_ and β_control_ are the coefficients for the weekly tournament-date fraction and matched-control-date fraction. Eight lags were selected to allow correlation across approximately two months of weekly observations; lags 4-12 were tested as sensitivity analyses. Because residual dependence remained substantial, adjusted models were used to evaluate robustness of the descriptive matched-window pattern.

### Sensitivity and Supplementary Analyses

Sensitivity analyses repeated the matched-window comparison using combined product-or-narrative, narrative-keyword-only, sports-location-restricted, age-stratified, tournament-plus-14-day, calendar-year, control-overlap, pandemic-exclusion, same-day-of-week-control, pre-2022-only, and leave-one-tournament-out definitions. We also compared the daily weighted percentage of the NEISS ED-treated consumer product/recreation injury population that was soccer-coded. Narrative-derived definitions were treated as exploratory.[21]

## RESULTS

### Cohort and Data Source

All 27 annual public NEISS files from 1999 through 2025 were analyzed, and all 9,794,932 records had valid treatment dates, weights, strata, and primary sampling units (Figure 1). The product-code case definition identified 170,679 soccer-coded records, corresponding to an estimated 5,366,681 ED-treated soccer-coded injuries nationally over the study period within the NEISS-estimated ED-treated consumer product and recreation-related injury population. No records were excluded by the plausibility screen. Annual field availability, including the 2018 addition of a third product field and later narrative-format changes, is shown in Supplementary Table S1. Mean daily weighted estimates were 453.1 injuries per day during tournament dates and 384.1 during matched control dates, corresponding to a difference of 68.9 injuries per day (95% CI, −0.5 to 138.3) and a ratio of 1.18 (95% CI, 1.00 to 1.39) (Table 1; Figure 2). Tournament-specific ratios ranged from 0.99 for the 2022 winter tournament to 1.31 in 2014 (Table 1; Figure 2).

**Figure 1.**
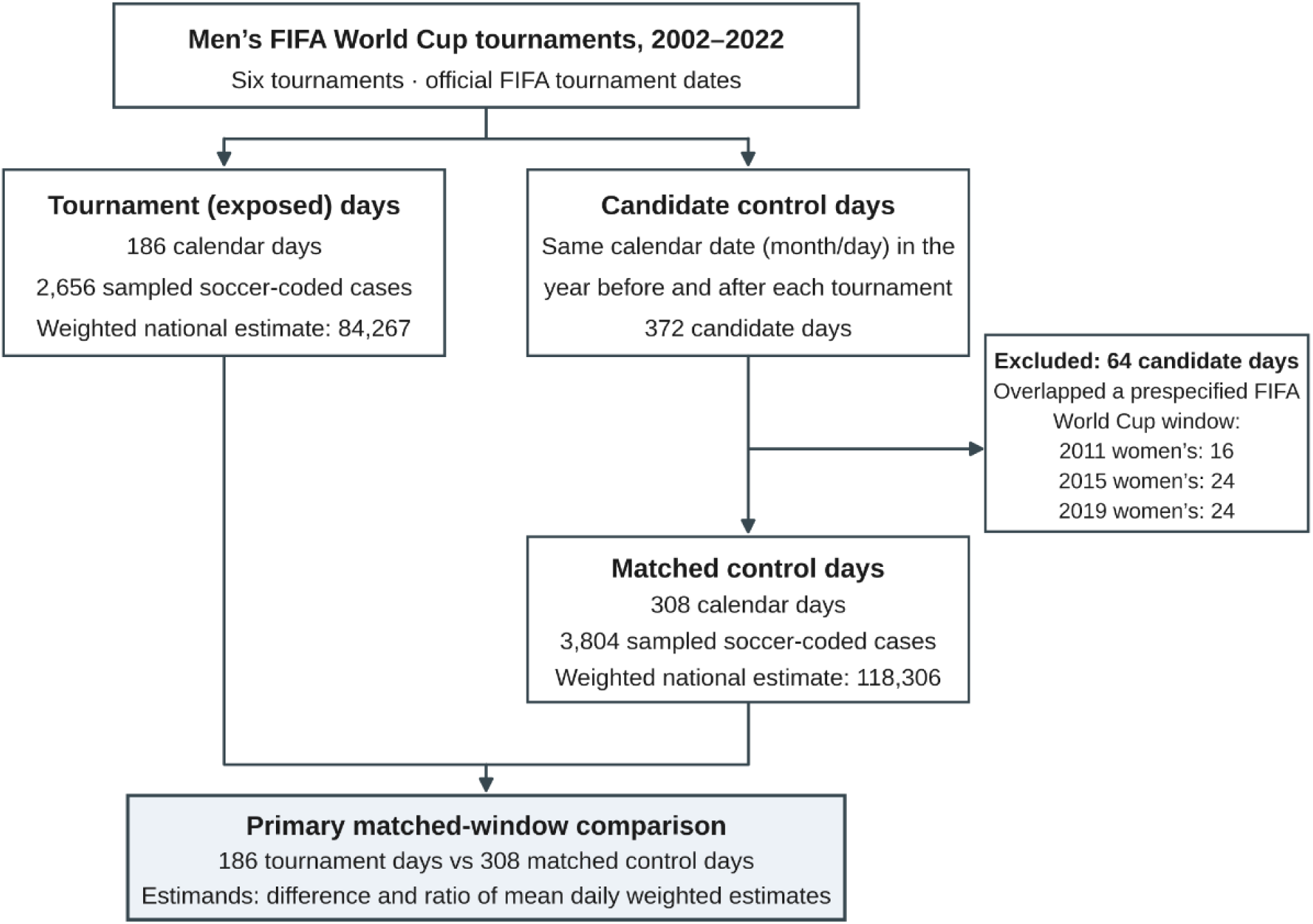
Study design schematic for the matched calendar-period surveillance analysis. The primary exposure comprised 186 official men’s FIFA World Cup tournament dates across six tournaments. Candidate controls were the same month/day in adjacent years (372 days); 64 candidate days overlapping prespecified men’s or women’s FIFA World Cup windows were excluded, leaving 308 matched control days. The primary estimands were the mean daily weighted estimate difference and ratio comparing tournament with matched-control dates.

**Table 1.**
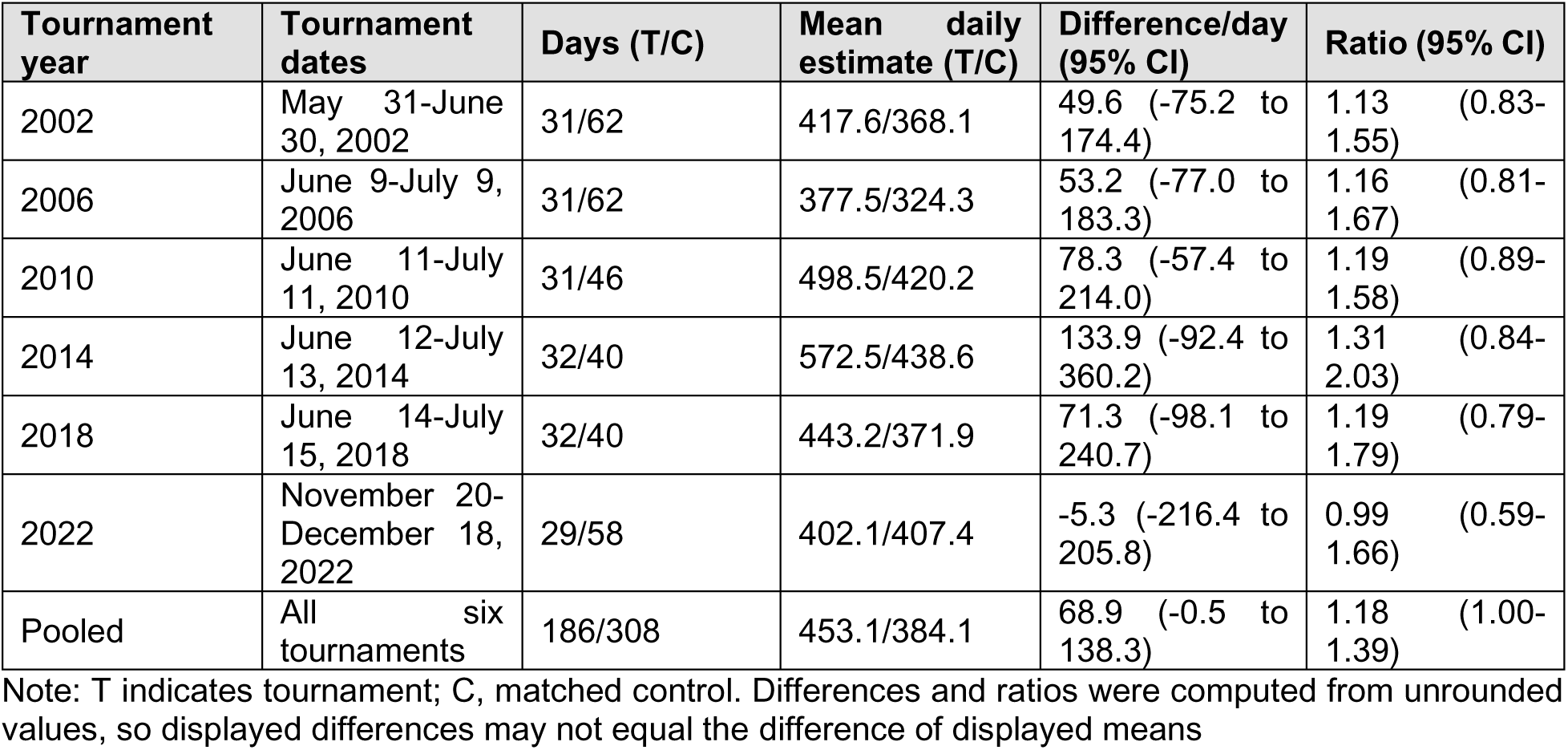
Tournament-specific and pooled matched-window comparisons of NEISS soccer-coded injuries, 1999–2025.

**Figure 2.**
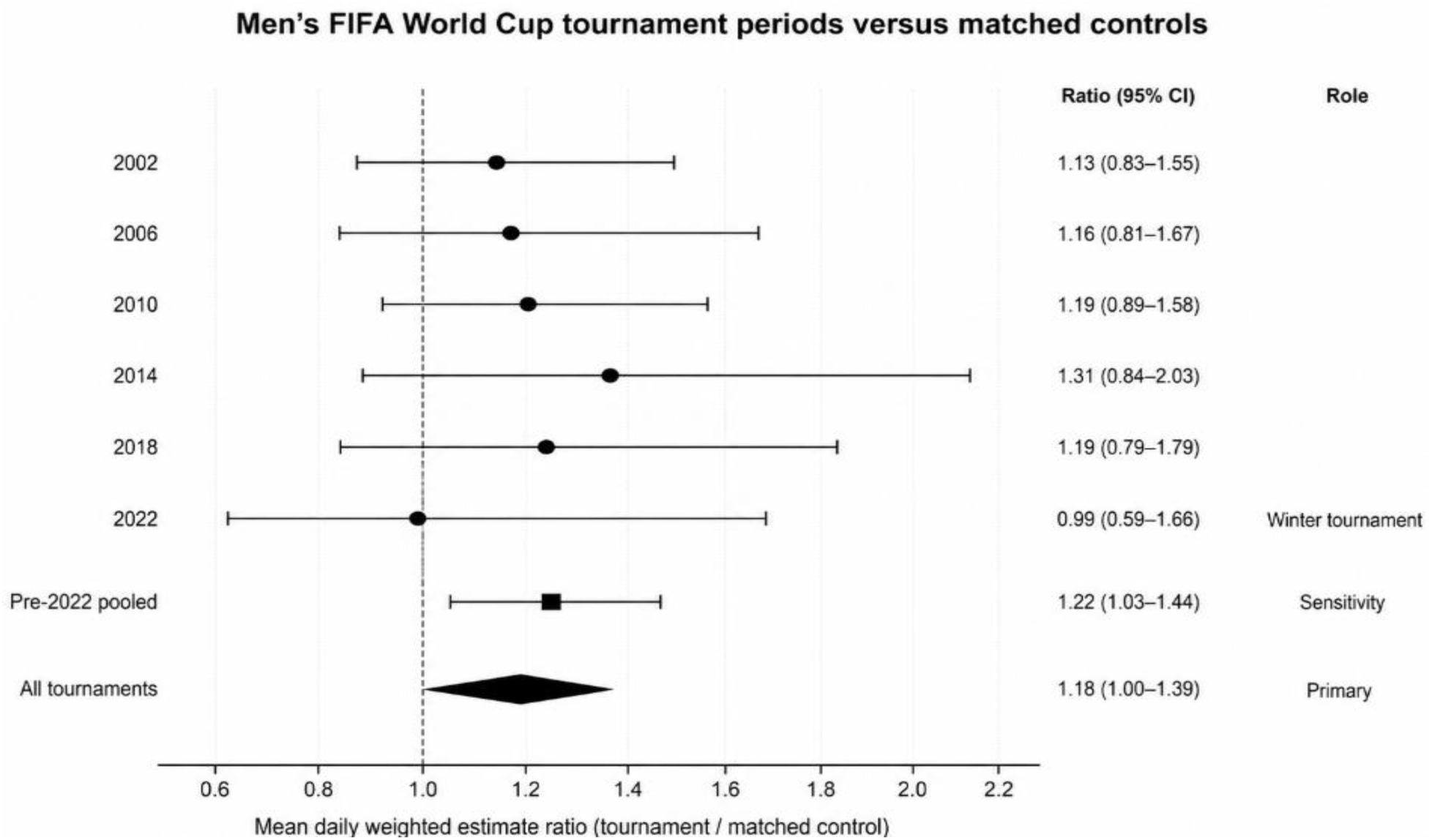
Tournament-specific and pooled mean daily weighted estimate ratios comparing men’s FIFA World Cup tournament periods with matched same-calendar control periods. Ratios use NEISS product code 1267 soccer-coded injuries; error bars show design-based 95% confidence intervals. The pooled primary estimate is shown with a diamond, the pre-2022 pooled estimate is shown as a sensitivity estimate, and the 2022 tournament is annotated as the winter tournament.

### Annual Product-Code-Defined Soccer Injury Burden

Annual weighted national estimates of ED-treated soccer-coded injuries ranged from 81,509 in 2020, during the COVID-19 pandemic, to 265,761 in 2024 (Supplementary Figure S1). Annual unweighted case counts ranged from 3,191 in 2020 to 8,951 in 2025. The weighted percentage of NEISS-estimated ED-treated consumer product and recreation-related injuries that were soccer-coded ranged from 0.74% in 2020 to 1.76% in 2024. Full annual estimates are provided in Supplementary Table S2.

### Men’s Tournament Periods Versus Matched Controls

During the 186 men’s tournament dates, 2,656 sampled cases yielded a weighted estimate of 84,267 ED-treated soccer-coded injuries, representing 1.15% of the NEISS ED-treated consumer product/recreation injury population in those periods. During the 308 matched same-calendar control dates, 3,804 sampled cases yielded a weighted estimate of 118,306, representing 0.99% of the corresponding denominator (Table 1). The paired percentage contrast was 0.16 percentage points (95% CI, 0.01 to 0.31), with a percentage ratio of 1.16 (95% CI, 1.01 to 1.33).

### Injury Characteristics Within the Soccer Cohort

Within the product-code-defined soccer cohort (170,679 records; weighted estimate, 5,366,681), the leading diagnoses were strain or sprain (31.2%), fracture (21.7%), and contusion or abrasion (15.0%); concussion accounted for 4.4%. The most frequently injured body parts were the ankle (17.0%), knee (12.6%), and head (11.2%). Most injuries occurred at sports or recreation places (62.6%) or schools (11.9%), and 97.2% of patients were treated or examined and released. Detailed injury-profile results are shown in Table 2.

**Table 2.** Diagnosis, body part, incident location, and disposition within the product-code-defined soccer cohort, NEISS 1999–2025. Denominator for all percentages: 170,679 unweighted records and 5,366,681 weighted ED-treated soccer-coded injuries. †Design-based 95% CIs. ‡Estimates with unweighted n < 20, weighted estimate < 1,200, or CV > 33% are suppressed as potentially unstable per CPSC screening criteria; unweighted counts are retained. National estimates were calculated using NEISS sample weights; unweighted counts represent sampled NEISS cases. Diagnosis and body-part categories shown are the leading stable categories.

| Domain | Category | Unweighted n | Weighted estimate | Weighted percent (95% CI) |
| --- | --- | --- | --- | --- |
| Diagnosis | Strain or sprain | 49,736 | 1,674,228 | 31.2 (30.6–31.8) |
| Diagnosis | Fracture | 40,321 | 1,164,676 | 21.7 (21.3–22.1) |
| Diagnosis | Contusion or abrasion | 23,619 | 807,112 | 15.0 (14.7–15.4) |
| Diagnosis | Other diagnosis | 23,482 | 706,995 | 13.2 (12.5–13.9) |
| Diagnosis | Internal injury | 9,577 | 281,864 | 5.3 (5.0–5.5) |
| Diagnosis | Laceration | 8,422 | 276,770 | 5.2 (5.0–5.3) |
| Diagnosis | Concussion | 8,564 | 234,343 | 4.4 (4.2–4.6) |
| Diagnosis | Dislocation | 4,324 | 144,316 | 2.7 (2.6–2.8) |
| Body part | Ankle | 27,266 | 910,174 | 17.0 (16.6–17.3) |
| Body part | Knee | 20,819 | 677,288 | 12.6 (12.4–12.9) |
| Body part | Head | 20,522 | 600,764 | 11.2 (10.9–11.5) |
| Body part | Wrist | 13,239 | 425,797 | 7.9 (7.7–8.1) |
| Body part | Foot | 9,249 | 310,346 | 5.8 (5.6–6.0) |
| Body part | Lower leg | 9,996 | 309,584 | 5.8 (5.6–5.9) |
| Body part | Face | 9,160 | 308,243 | 5.7 (5.6–5.9) |
| Body part | Finger | 9,311 | 293,266 | 5.5 (5.3–5.6) |
| Body part | Shoulder | 8,037 | 257,287 | 4.8 (4.6–4.9) |
| Location | Sports or recreation place | 113,375 | 3,358,537 | 62.6 (60.4–64.7) |
| Location | Unknown or not stated | 33,068 | 1,138,014 | 21.2 (19–23.4) |
| Location | School | 17,734 | 639,652 | 11.9 (11.2–12.6) |
| Location | Home | 4,940 | 167,338 | 3.1 (2.9–3.3) |
| Location | Other public property | 1,239 | 53,118 | 1.0 (0.8–1.2) |
| Location | Street or highway | 309 | 9,586 | 0.18 (0.14–0.22) |
| Location | Mobile or manufactured home | 8 | Suppressed‡ | Suppressed‡ |
| Location | Farm or ranch | 4 | Suppressed<sup>‡</sup> | Suppressed<sup>‡</sup> |
| Location | Industrial place | 2 | Suppressed<sup>‡</sup> | Suppressed<sup>‡</sup> |
| Disposition | Treated or examined and released | 164,927 | 5,213,770 | 97.2 (97–97.3) |
| Disposition | Treated and admitted or hospitalized | 3,576 | 84,583 | 1.6 (1.5–1.7) |
| Disposition | Left without being seen | 1,361 | 40,986 | 0.76 (0.68–0.85) |
| Disposition | Treated and transferred | 398 | 19,881 | 0.37 (0.32–0.42) |
| Disposition | Held for observation | 375 | 6,252 | 0.12 (0.09–0.14) |
| Disposition | Unknown or not stated | 25 | Suppressed<sup>‡</sup> | Suppressed<sup>‡</sup> |
| Disposition | Fatality, including dead on arrival | 17 | Suppressed<sup>‡</sup> | Suppressed<sup>‡</sup> |

### Model Sensitivity Analyses

Across the 1,408 analyzed weeks, raw model residuals showed strong positive autocorrelation, and the quasi-Poisson specification indicated overdispersion. Accordingly, adjusted models were interpreted as secondary robustness checks rather than primary evidence. The tournament-versus-control adjusted ratio was 1.18 (95% CI, 1.01 to 1.38) in the survey-weighted AR(1) WLS/HAC model, 1.15 (95% CI, 0.99 to 1.34) in the non-autoregressive WLS/HAC model, 1.15 (95% CI, 0.99 to 1.34) in the calendar-week fixed-effect WLS/HAC model, and 1.15 (95% CI, 1.01 to 1.31) in the quasi-Poisson/HAC model (Supplementary Table S5). Newey-West lag sensitivity showed the same point estimate at 4 to 12 lags, with CIs ranging from 0.99 to 1.40 at 4 lags to 1.02 to 1.37 at 12 lags. The AR(1) parameter was 0.63 in the primary WLS/HAC model, and the quasi-Poisson/HAC sensitivity had Pearson dispersion 5.84.

### Outcome-Definition, Window, and Subgroup Sensitivity Analyses

Sensitivity analyses are summarized in Supplementary Table S6. The same-day-of-week matched-control sensitivity yielded a ratio of 1.17 (95% CI, 0.99 to 1.37), similar in magnitude to the primary 1.18 ratio but with an interval crossing the null. The pre-2022-only sensitivity yielded a ratio of 1.22 (95% CI, 1.03 to 1.44), whereas the 2022 tournament-specific ratio was 0.99, indicating seasonal and tournament-context variation rather than a uniform World Cup association. Additional sensitivity estimates were consistent in direction but not uniform: product-or-narrative definition ratio 1.18 (95% CI, 1.00 to 1.38), narrative-keyword-only ratio 1.17 (95% CI, 0.99 to 1.38), sports-location-restricted ratio 1.18 (95% CI, 0.96 to 1.44), tournament-plus-14-day ratio 1.19 (95% CI, 1.02 to 1.39), calendar-year ratio 1.01 (95% CI, 0.90 to 1.14), and pandemic-exclusion ratio 1.17 (95% CI, 1.00 to 1.38). Age-stratified ratios were 1.12 for ages 0-9 years, 1.15 for 10-19 years, 1.23 for 20-39 years, and 1.29 for 40 years or older.

## DISCUSSION

### Principal Findings

In this national surveillance analysis of ED-treated soccer-coded injuries, men’s FIFA World Cup tournament dates had a modestly higher mean daily NEISS-estimated injury count than matched adjacent-year same-calendar dates, but uncertainty was substantial and tournament-specific results were heterogeneous. The 95% CI for the ratio reached 1.00 after rounding, and the CI for the absolute difference included zero. These findings support surveillance preparedness for major soccer events but do not support inference about individual-level World Cup exposure.

### Interpretation of Window-Level and Calendar-Year Estimates

The tournament-window ratio (1.18) contrasted with the near-null calendar-year ratio (1.01), illustrating how classifying an entire year as exposed can dilute a short-term calendar-period pattern.[11] This contrast supports the event-window design for surveillance questions. The adjusted weekly models addressed a related but distinct question after accounting for trend, seasonality, and the pandemic period.

The 2022 result is particularly informative because it occurred during late autumn and early winter in the United States, unlike the summer tournaments from 2002 through 2018. The winter timing may have reduced outdoor recreational soccer opportunity or changed school, weather, daylight, and participation patterns. Broadcast timing may also have modified viewing behavior across tournaments; for example, tournaments outside U.S. time zones may have shifted viewing away from typical recreational play times. Conversely, intensive viewing could displace play and attenuate an injury signal. These competing mechanisms support interpreting the findings as heterogeneous calendar-period surveillance signals rather than as evidence of a direct tournament-to-injury pathway.

### Comparison With Prior Literature

The injury profile observed within the soccer cohort, dominated by sprains and strains, fractures, and contusions, with the ankle, knee, and head as leading body parts, is consistent with prior NEISS-based soccer studies in children and young adults.[22, 23] Event-window studies around World Cup tournaments have reported short-term changes in cardiovascular admissions during match periods in viewing populations,[7] and standardized FIFA surveillance has characterized injuries among tournament players.[9] However, neither body of literature has examined population-level ED-treated soccer-coded injuries.[9]

### Public Health Implications

The observed pattern may help guide injury surveillance around major soccer events and inform targeted prevention activities. Practical applications include near-real-time syndromic surveillance systems could monitor soccer-related ED presentations during tournament periods, youth and adult recreational soccer injury-prevention messaging, concussion recognition and return-to-play messaging,[24] monitoring of fan-zone and community-event injury patterns, and linkage of ED data with urgent care, athletic trainer, emergency medical services, weather, and participation data when feasible.[25, 26] Future studies could link ED injury data with participation, viewership, local tournament engagement, and community events.[27]

For 2026 North American World Cup planning, the practical implication is surveillance readiness.[28] ED surveillance teams and public health partners could prespecify soccer-coded injury monitoring, timely review of narrative text, youth and adult injury-prevention messaging, and rapid contextual assessment of participation or fan-event activity during tournament weeks.

### Strengths and Limitations

This study has several strengths. NEISS is a nationally representative probability-sample surveillance system, allowing survey-weighted estimates with design-based uncertainty. The 27-year study period included six men’s tournaments and matched calendar controls, with analyses accounting for secular trend, seasonality, and pandemic disruption. Adjusted models and alternative outcome definitions were used as sensitivity analyses. Structured product codes defined the primary cohort, and narrative information was limited to supplementary analyses to avoid overinterpreting brief and documentation-dependent free text.

Several limitations should be considered. First, NEISS represents injuries treated in participating hospital EDs; injuries treated in athletic training rooms, urgent care centers, outpatient clinics, or not treated medically are not captured, so the estimates describe ED-treated burden rather than total soccer injury occurrence, and participation denominators were unavailable.[3] Second, NEISS product coding identifies product- or activity-associated injuries and does not establish causality, and World Cup indicators are ecological calendar-time variables that do not establish individual exposure to playing, attending, or viewing tournaments. Third, Product 3 became available only in 2018, which may have slightly increased soccer-code ascertainment after that date and could affect the 2018 tournament/control comparison. Fourth, residual autocorrelation and model-specification sensitivity limit adjusted model interpretation. Fifth, this study did not include negative-control sports, women’s World Cup exposure windows,[29] placebo windows, or formal minimum detectable effect calculations; these analyses could further assess specificity and precision in future work.[30]

## CONCLUSIONS

In this national analysis of NEISS data, mean daily estimates of ED-treated soccer-coded injuries were modestly higher during men’s FIFA World Cup tournament periods than during matched control periods. Their principal value is practical: major soccer tournaments are predictable, well-defined windows in which ED-based injury surveillance, recreational soccer injury-prevention and concussion-awareness messaging, and monitoring of community and fan-event injury patterns can be organized in advance. The 2026 FIFA World Cup in North America, and each tournament cycle thereafter, offers a recurring opportunity to prespecify such surveillance and test the patterns.

## Declarations

### Ethics approval and consent to participate

This study used publicly available, de-identified NEISS data and did not constitute human subjects research; institutional review board approval and informed consent were not required.

### Consent for publication

Not applicable.

### Availability of data and materials

All data analyzed during this study are available at the National Electronic Injury Surveillance System: https://www.cpsc.gov/Research--Statistics/NEISS-Injury-Data; code and supporting materials are available at: https://github.com/rspraneethk2076/NEISS_Analysis/tree/NEISS_Analysis_World_cup.

### Authors’ contributions

JY designed the study. JY and RSPK performed the data extraction and statistical analysis and drafted the manuscript. All authors contributed to interpretation of the results, critical revision of the manuscript for important intellectual content, and approved the final version for submission.

## Data Availability

All data analyzed during this study are available at the National Electronic Injury Surveillance System: https://www.cpsc.gov/Research--Statistics/NEISS-Injury-Data

## Acknowledgments

None.

## Supplementary Material

**Supplementary Table S1.** Annual NEISS file availability, field mapping, and soccer-coded record counts.

| Year | Database records | Eligible records | Treatment dates | Product fields | Narrative fields | Soccer-coded n | Soccer-coded weighted estimate |
| --- | --- | --- | --- | --- | --- | --- | --- |
| 1999 | 312,943 | 312,943 | 1999-01-01 to 1999-12-31 | P1/P2 | N1/N2 | 4,540 | 175,303 |
| 2000 | 339,244 | 339,244 | 2000-01-01 to 2000-12-31 | P1/P2 | N1/N2 | 4,857 | 185,064 |
| 2001 | 356,038 | 356,038 | 2001-01-01 to 2001-12-31 | P1/P2 | N1/N2 | 4,843 | 175,470 |
| 2002 | 359,980 | 359,980 | 2002-01-01 to 2002-12-31 | P1/P2 | N1/N2 | 4,967 | 173,519 |
| 2003 | 347,380 | 347,380 | 2003-01-01 to 2003-12-31 | P1/P2 | N1/N2 | 4,648 | 166,791 |
| 2004 | 353,394 | 353,394 | 2004-01-01 to 2004-12-31 | P1/P2 | N1/N2 | 4,908 | 173,509 |
| 2005 | 360,374 | 360,374 | 2005-01-01 to 2005-12-31 | P1/P2 | N1/N2 | 5,285 | 174,686 |
| 2006 | 363,616 | 363,616 | 2006-01-01 to 2006-12-31 | P1/P2 | N1/N2 | 5,517 | 186,544 |
| 2007 | 369,841 | 369,841 | 2007-01-01 to 2007-12-31 | P1/P2 | N1/N2 | 6,003 | 198,679 |
| 2008 | 374,260 | 374,260 | 2008-01-01 to 2008-12-31 | P1/P2 | N1/N2 | 6,187 | 199,475 |
| 2009 | 391,944 | 391,944 | 2009-01-01 to 2009-12-31 | P1/P2 | N1/N2 | 6,628 | 208,214 |
| 2010 | 405,710 | 405,710 | 2010-01-01 to 2010-12-31 | P1/P2 | N1/N2 | 7,250 | 226,142 |
| 2011 | 396,502 | 396,502 | 2011-01-01 to 2011-12-31 | P1/P2 | N1/N2 | 6,985 | 214,053 |
| 2012 | 394,383 | 394,383 | 2012-01-01 to 2012-12-31 | P1/P2 | N1/N2 | 7,347 | 231,447 |
| 2013 | 376,926 | 376,926 | 2013-01-01 to 2013-12-31 | P1/P2 | N1/N2 | 7,274 | 229,088 |
| 2014 | 367,493 | 367,493 | 2014-01-01 to 2014-12-31 | P1/P2 | N1/N2 | 7,574 | 239,943 |
| 2015 | 359,129 | 359,129 | 2015-01-01 to 2015-12-31 | P1/P2 | N1/N2 | 7,245 | 227,732 |
| 2016 | 375,197 | 375,197 | 2016-01-01 to 2016-12-31 | P1/P2 | N1/N2 | 7,671 | 226,595 |
| 2017 | 386,906 | 386,906 | 2017-01-01 to 2017-12-31 | P1/P2 | N1/N2 | 7,550 | 218,926 |
| 2018 | 361,667 | 361,667 | 2018-01-01 to 2018-12-31 | P1/P2/P3 | N1 | 6,789 | 199,096 |
| 2019 | 358,715 | 358,715 | 2019-01-01 to 2019-12-31 | P1/P2/P3 | N1 | 6,591 | 188,336 |
| 2020 | 309,370 | 309,370 | 2020-01-01 to 2020-12-31 | P1/P2/P3 | N1 | 3,191 | 81,509 |
| 2021 | 340,442 | 340,442 | 2021-01-01 to 2021-12-31 | P1/P2/P3 | N1 | 5,611 | 144,895 |
| 2022 | 323,343 | 323,343 | 2022-01-01 to 2022-12-31 | P1/P2/P3 | N1 | 6,308 | 179,284 |
| 2023 | 338,262 | 338,262 | 2023-01-01 to 2023-12-31 | P1/P2/P3 | N1 | 7,603 | 212,423 |
| 2024 | 361,672 | 361,672 | 2024-01-01 to 2024-12-31 | P1/P2/P3 | N1 | 8,356 | 265,761 |
| 2025 | 410,201 | 410,201 | 2025-01-01 to 2025-12-31 | P1/P2/P3 | N1 | 8,951 | 264,199 |

Note: P1-P3 indicate available product-code fields; N1-N2 indicate available narrative fields. Soccer-coded injuries were identified using NEISS product code 1267 in any available product field.

**Supplementary Table S2.** Annual soccer-coded injury burden and denominator estimates, NEISS 1999-2025.

| Year | Men's World Cup year | Unweighted n | Weighted estimate | 95% CI | Weighted % | Denominator weighted estimate |
| --- | --- | --- | --- | --- | --- | --- |
| 1999 | No | 4,540 | 175,303 | 136,246 to 214,361 | 1.44% | 12,165,225 |
| 2000 | No | 4,857 | 185,064 | 136,191 to 233,937 | 1.43% | 12,924,488 |
| 2001 | No | 4,843 | 175,470 | 142,384 to 208,555 | 1.32% | 13,310,088 |
| 2002 | Yes | 4,967 | 173,519 | 138,834 to 208,204 | 1.36% | 12,790,786 |
| 2003 | No | 4,648 | 166,791 | 129,025 to 204,557 | 1.31% | 12,720,947 |
| 2004 | No | 4,908 | 173,509 | 132,455 to 214,562 | 1.32% | 13,096,983 |
| 2005 | No | 5,285 | 174,686 | 133,350 to 216,023 | 1.39% | 12,608,693 |
| 2006 | Yes | 5,517 | 186,544 | 142,183 to 230,904 | 1.41% | 13,232,263 |
| 2007 | No | 6,003 | 198,679 | 151,187 to 246,172 | 1.50% | 13,232,338 |
| 2008 | No | 6,187 | 199,475 | 151,668 to 247,282 | 1.48% | 13,456,353 |
| 2009 | No | 6,628 | 208,214 | 162,719 to 253,708 | 1.49% | 13,966,898 |
| 2010 | Yes | 7,250 | 226,142 | 178,066 to 274,219 | 1.54% | 14,694,928 |
| 2011 | No | 6,985 | 214,053 | 166,055 to 262,052 | 1.51% | 14,162,084 |
| 2012 | No | 7,347 | 231,447 | 177,135 to 285,759 | 1.58% | 14,614,128 |
| 2013 | No | 7,274 | 229,088 | 170,418 to 287,759 | 1.63% | 14,033,745 |
| 2014 | Yes | 7,574 | 239,943 | 171,297 to 308,589 | 1.73% | 13,860,971 |
| 2015 | No | 7,245 | 227,732 | 160,870 to 294,593 | 1.61% | 14,132,697 |
| 2016 | No | 7,671 | 226,595 | 153,399 to 299,790 | 1.58% | 14,319,418 |
| 2017 | No | 7,550 | 218,926 | 158,344 to 279,507 | 1.49% | 14,727,704 |
| 2018 | Yes | 6,789 | 199,096 | 142,805 to 255,386 | 1.43% | 13,959,174 |
| 2019 | No | 6,591 | 188,336 | 132,739 to 243,934 | 1.40% | 13,464,372 |
| 2020 | No | 3,191 | 81,509 | 52,977 to 110,040 | 0.74% | 10,994,077 |
| 2021 | No | 5,611 | 144,895 | 105,454 to 184,335 | 1.23% | 11,738,091 |
| 2022 | Yes | 6,308 | 179,284 | 126,345 to 232,223 | 1.42% | 12,663,028 |
| 2023 | No | 7,603 | 212,423 | 154,068 to 270,777 | 1.67% | 12,740,256 |
| 2024 | No | 8,356 | 265,761 | 207,779 to 323,744 | 1.76% | 15,068,979 |
| 2025 | No | 8,951 | 264,199 | 199,442 to 328,955 | 1.60% | 16,485,413 |

Note: Numerator is NEISS product code 1267 in any available product field. Denominator is NEISS-estimated emergency department-treated consumer product and recreation-related injuries.

**Supplementary Table S3.**
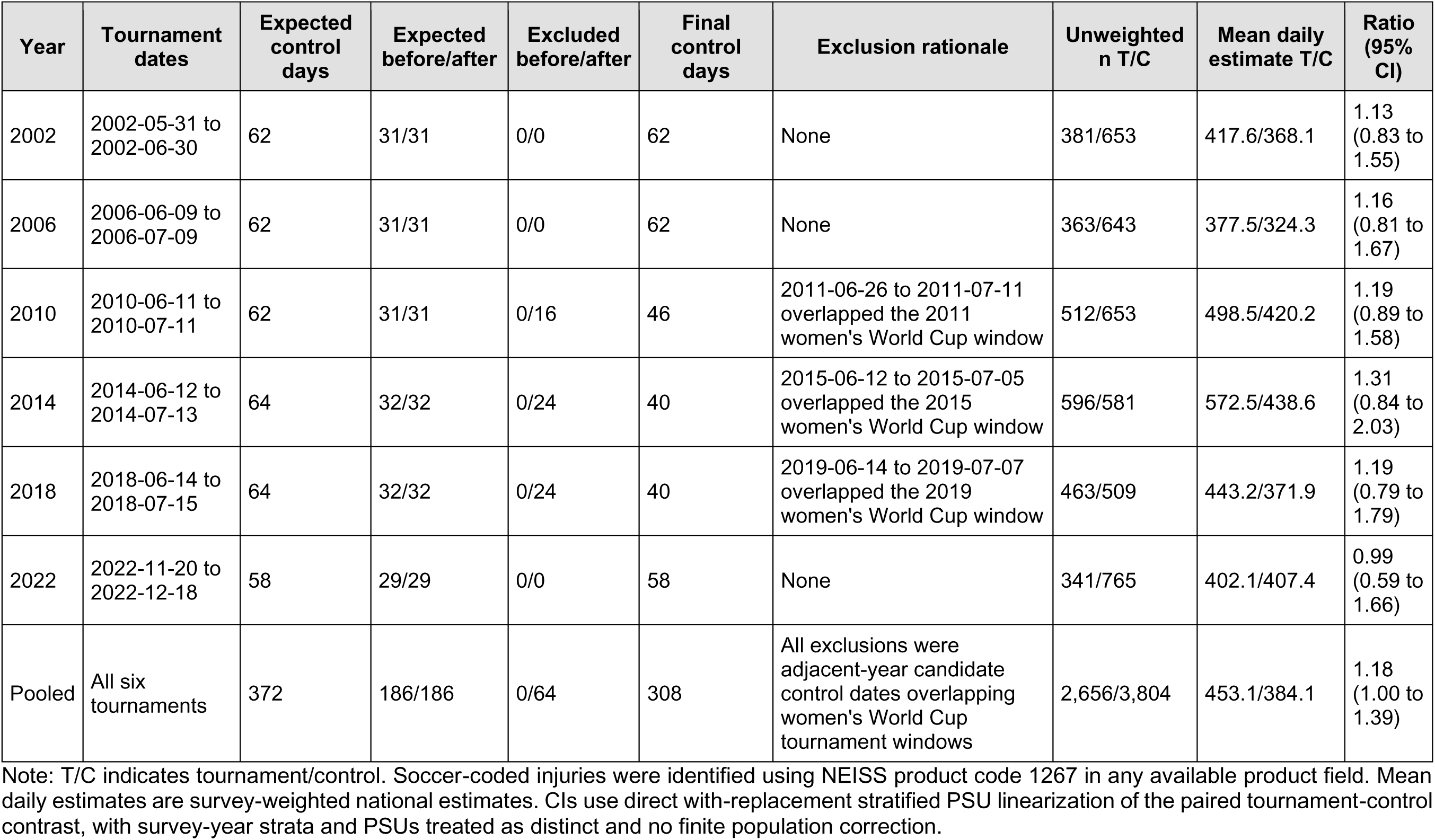
FIFA tournament calendar and matched-control exclusion audit.

| Year | Tournament dates | Expected control days | Expected before/after | Excluded before/after | Final control days | Exclusion rationale | Unweighted n T/C | Mean daily estimate T/C | Ratio (95% CI) |
| --- | --- | --- | --- | --- | --- | --- | --- | --- | --- |
| 2002 | 2002-05-31 to 2002-06-30 | 62 | 31/31 | 0/0 | 62 | None | 381/653 | 417.6/368.1 | 1.13 (0.83 to 1.55) |
| 2006 | 2006-06-09 to 2006-07-09 | 62 | 31/31 | 0/0 | 62 | None | 363/643 | 377.5/324.3 | 1.16 (0.81 to 1.67) |
| 2010 | 2010-06-11 to 2010-07-11 | 62 | 31/31 | 0/16 | 46 | 2011-06-26 to 2011-07-11 overlapped the 2011 women's World Cup window | 512/653 | 498.5/420.2 | 1.19 (0.89 to 1.58) |
| 2014 | 2014-06-12 to 2014-07-13 | 64 | 32/32 | 0/24 | 40 | 2015-06-12 to 2015-07-05 overlapped the 2015 women's World Cup window | 596/581 | 572.5/438.6 | 1.31 (0.84 to 2.03) |
| 2018 | 2018-06-14 to 2018-07-15 | 64 | 32/32 | 0/24 | 40 | 2019-06-14 to 2019-07-07 overlapped the 2019 women's World Cup window | 463/509 | 443.2/371.9 | 1.19 (0.79 to 1.79) |
| 2022 | 2022-11-20 to 2022-12-18 | 58 | 29/29 | 0/0 | 58 | None | 341/765 | 402.1/407.4 | 0.99 (0.59 to 1.66) |
| Pooled | All six tournaments | 372 | 186/186 | 0/64 | 308 | All exclusions were adjacent-year candidate control dates overlapping women's World Cup tournament windows | 2,656/3,804 | 453.1/384.1 | 1.18 (1.00 to 1.39) |

Note: T/C indicates tournament/control. Soccer-coded injuries were identified using NEISS product code 1267 in any available product field. Mean daily estimates are survey-weighted national estimates. CIs use direct with-replacement stratified PSU linearization of the paired tournament-control contrast, with survey-year strata and PSUs treated as distinct and no finite population correction.

**Supplementary Table S4.** Primary and closely related matched-window contrast outputs.

| Analysis | Outcome | Days T/C | Unweighted<br>n T/C | Weighted<br>estimate T/C | Mean daily<br>T/C | Difference<br>(95% CI) | Ratio (95%<br>CI) | Variance method |
| --- | --- | --- | --- | --- | --- | --- | --- | --- |
| Primary<br>direct paired<br>PSU<br>contrast | soccer_product | 186/308 | 2,656/3,804 | 84,267/118,306 | 453.1/384.1 | 68.9 (-0.5 to<br>138.3) | 1.18 (1.00<br>to 1.39) | Direct with-replacement stratified<br>PSU linearization of<br>the paired contrast;<br>survey-year strata and<br>PSUs treated as<br>distinct; no finite<br>population correction. |
| Same-day-of-week<br>matched<br>control<br>sensitivity | soccer_product | 186/307 | 2,656/3,824 | 84,267/119,294 | 453.1/388.6 | 64.5 (-5.1 to<br>134.0) | 1.17 (0.99<br>to 1.37) | Direct with-replacement stratified<br>PSU linearization of<br>the paired contrast;<br>survey-year strata and<br>PSUs treated as<br>distinct; no finite<br>population correction. |
| Pre-2022-only<br>sensitivity | soccer_product | 157/250 | 2,315/3,039 | 72,606/94,677 | 462.5/378.7 | 83.8 (12.4<br>to 155.1) | 1.22 (1.03<br>to 1.44) | Direct with-replacement stratified<br>PSU linearization of<br>the paired contrast;<br>survey-year strata and<br>PSUs treated as<br>distinct; no finite<br>population correction. |

Note: T/C indicates tournament/control. Weighted estimates use NEISS sample weights; ratios compare mean daily weighted estimates.

**Supplementary Table S5.**
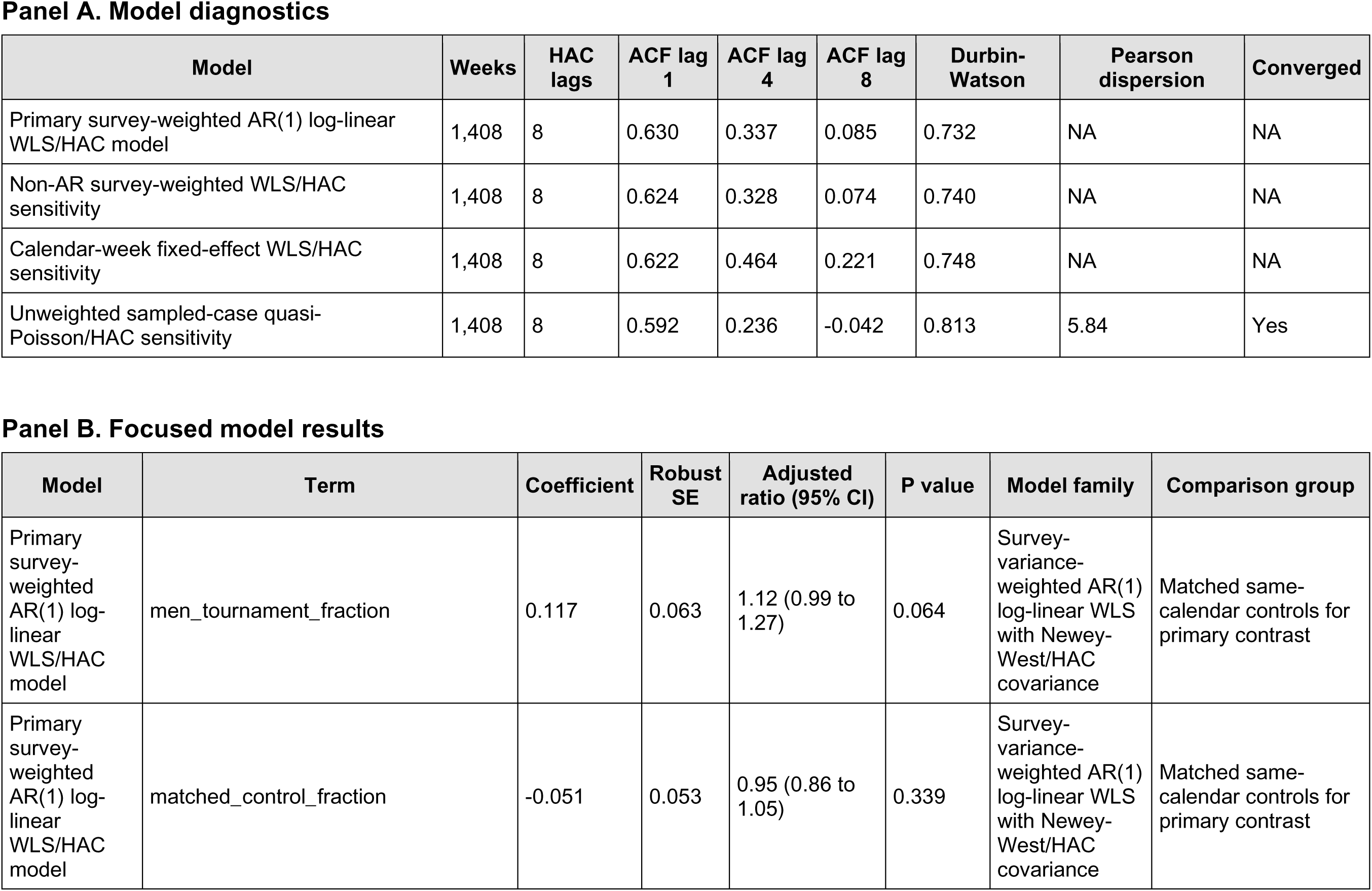
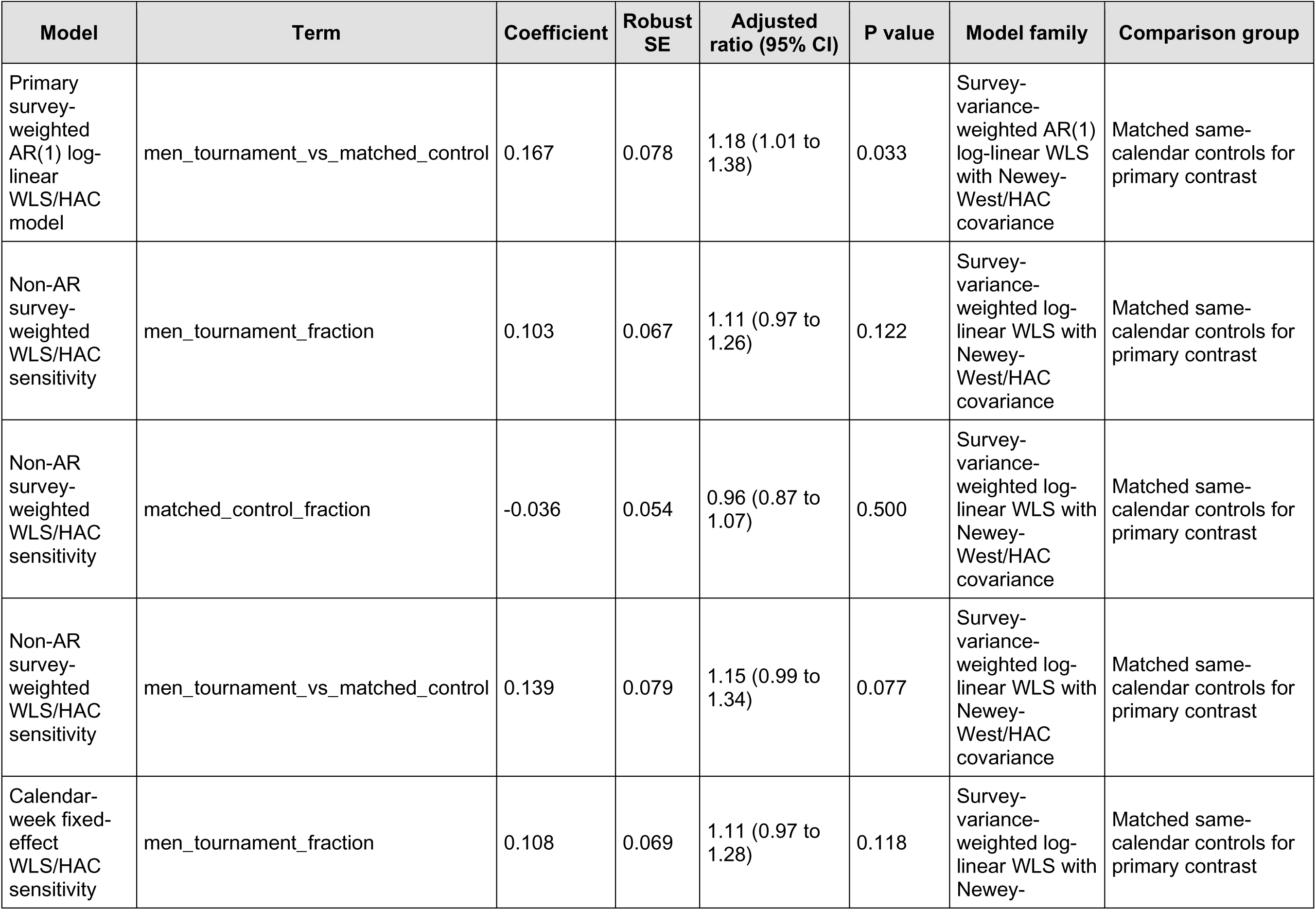
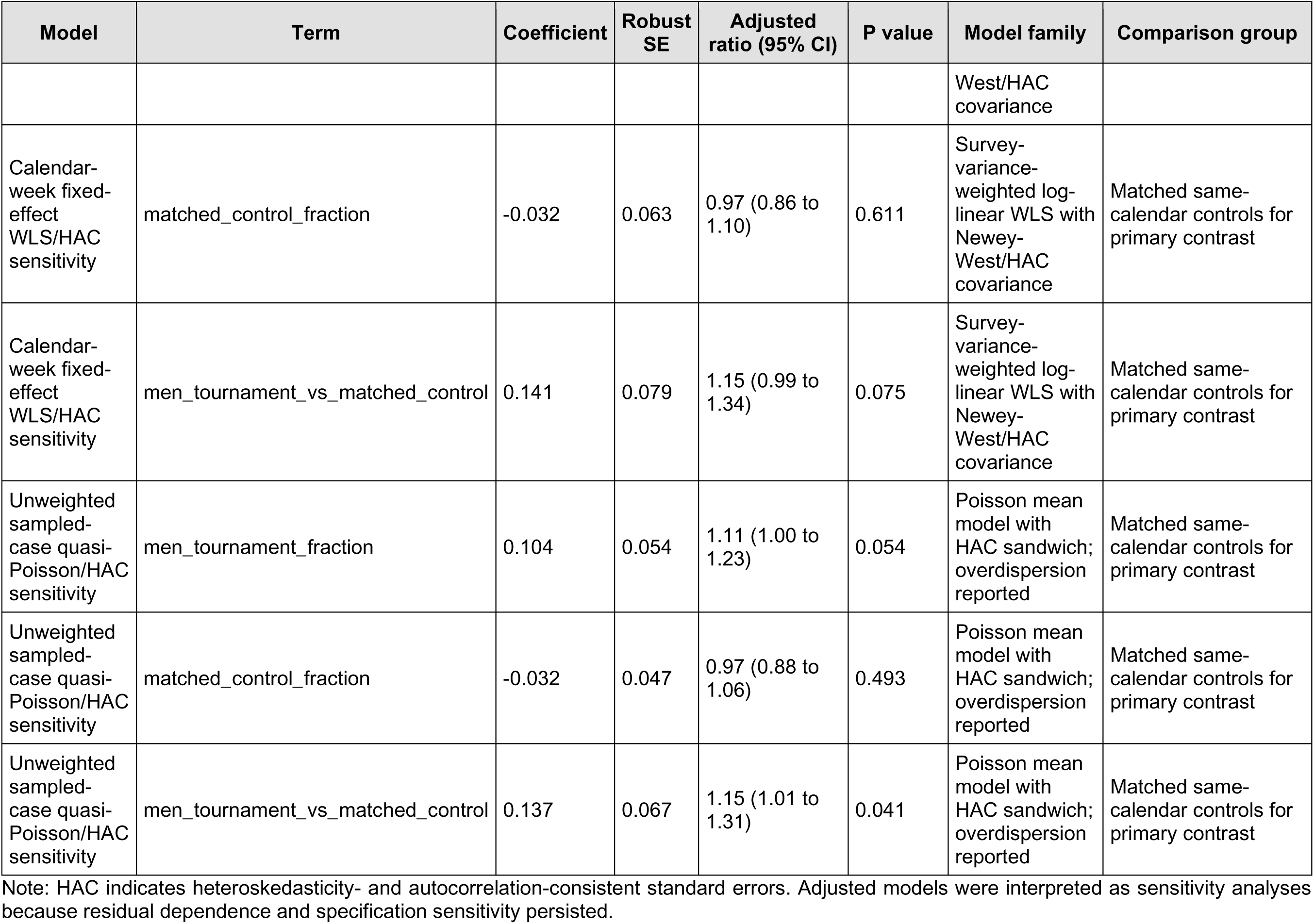
Weekly model diagnostics and focused model results. Panel A. Model diagnostics.

**Supplementary Table S6.** Outcome-definition, window, subgroup, and robustness sensitivity analyses.

| Outcome | Days T/C | Unweighted n T/C | Mean daily T/C | Difference (95% CI) | Ratio (95% CI) | Sensitivity definition |
| --- | --- | --- | --- | --- | --- | --- |
| soccer_product | 186/308 | 2,656/3,804 | 453.1/384.1 | 68.9 (-0.5 to 138.3) | 1.18 (1.00 to 1.39) | soccer_product |
| soccer_combined | 186/308 | 2,670/3,831 | 454.0/386.1 | 67.9 (-1.7 to 137.5) | 1.18 (1.00 to 1.38) | soccer_combined |
| soccer_narrative | 186/308 | 2,627/3,783 | 442.2/379.1 | 63.1 (-6.0 to 132.1) | 1.17 (0.99 to 1.38) | soccer_narrative |
| soccer_sports_location | 186/308 | 1,877/2,664 | 301.2/256.3 | 44.9 (-13.8 to 103.5) | 1.18 (0.96 to 1.44) | soccer_sports_location |
| soccer_age_0_9 | 186/308 | 336/477 | 46.5/41.6 | 4.8 (-7.6 to 17.3) | 1.12 (0.85 to 1.47) | soccer_age_0_9 |
| soccer_age_10_19 | 186/308 | 1,447/2,053 | 232.3/201.5 | 30.7 (-11.8 to 73.3) | 1.15 (0.95 to 1.40) | soccer_age_10_19 |
| soccer_age_20_39 | 186/308 | 730/1,066 | 144.0/117.5 | 26.6 (2.7 to 50.5) | 1.23 (1.02 to 1.47) | soccer_age_20_39 |
| soccer_age_40_plus | 186/308 | 143/208 | 30.2/23.4 | 6.8 (-1.3 to 14.9) | 1.29 (0.96 to 1.73) | soccer_age_40_plus |
| soccer_product | 270/384 | 3,784/4,630 | 440.5/371.2 | 69.3 (5.2 to 133.4) | 1.19 (1.02 to 1.39) | soccer_product; tournament plus 14 days |
| soccer_product | 2,190/7,672 | 38,405/132,274 | 550.0/542.5 | 7.5 (-58.1 to 73.1) | 1.01 (0.90 to 1.14) | soccer_product; men's World Cup calendar year |
| soccer_product | 186/279 | 2,656/3,439 | 453.1/386.1 | 67.0 (-3.0 to 136.9) | 1.17 (1.00 to 1.38) | soccer_product; primary windows excluding 2020-2021 |

Note: T/C indicates tournament/control. Ratios compare mean daily weighted national estimates between tournament and matched-control dates. — and consider merging the redundant Outcome/Sensitivity-definition columns

**Supplementary Table S7.** CPSC NEISS Query Builder calibration for soccer product code 1267, 2021-2025.

| Year | CPSC unweighted cases | Local unweighted n | Unweighted match | CPSC weighted estimate | Local weighted estimate | Relative difference (%) | Estimate match after rounding |
| --- | --- | --- | --- | --- | --- | --- | --- |
| 2021 | 5,611 | 5,611 | Yes | 144,895 | 144,895 | 0.00 | Yes |
| 2022 | 6,308 | 6,308 | Yes | 179,284 | 179,284 | 0.00 | Yes |
| 2023 | 7,603 | 7,603 | Yes | 212,423 | 212,423 | 0.00 | Yes |
| 2024 | 8,356 | 8,356 | Yes | 265,761 | 265,761 | 0.00 | Yes |
| 2025 | 8,951 | 8,951 | Yes | 264,199 | 264,199 | 0.00 | Yes |

Note: Calibration compared local processing against the official CPSC NEISS Query Builder for soccer product code 1267. Confidence-limit endpoints may differ because public-use variance implementation details are not identical to CPSC output.

**Supplementary Table S8.**
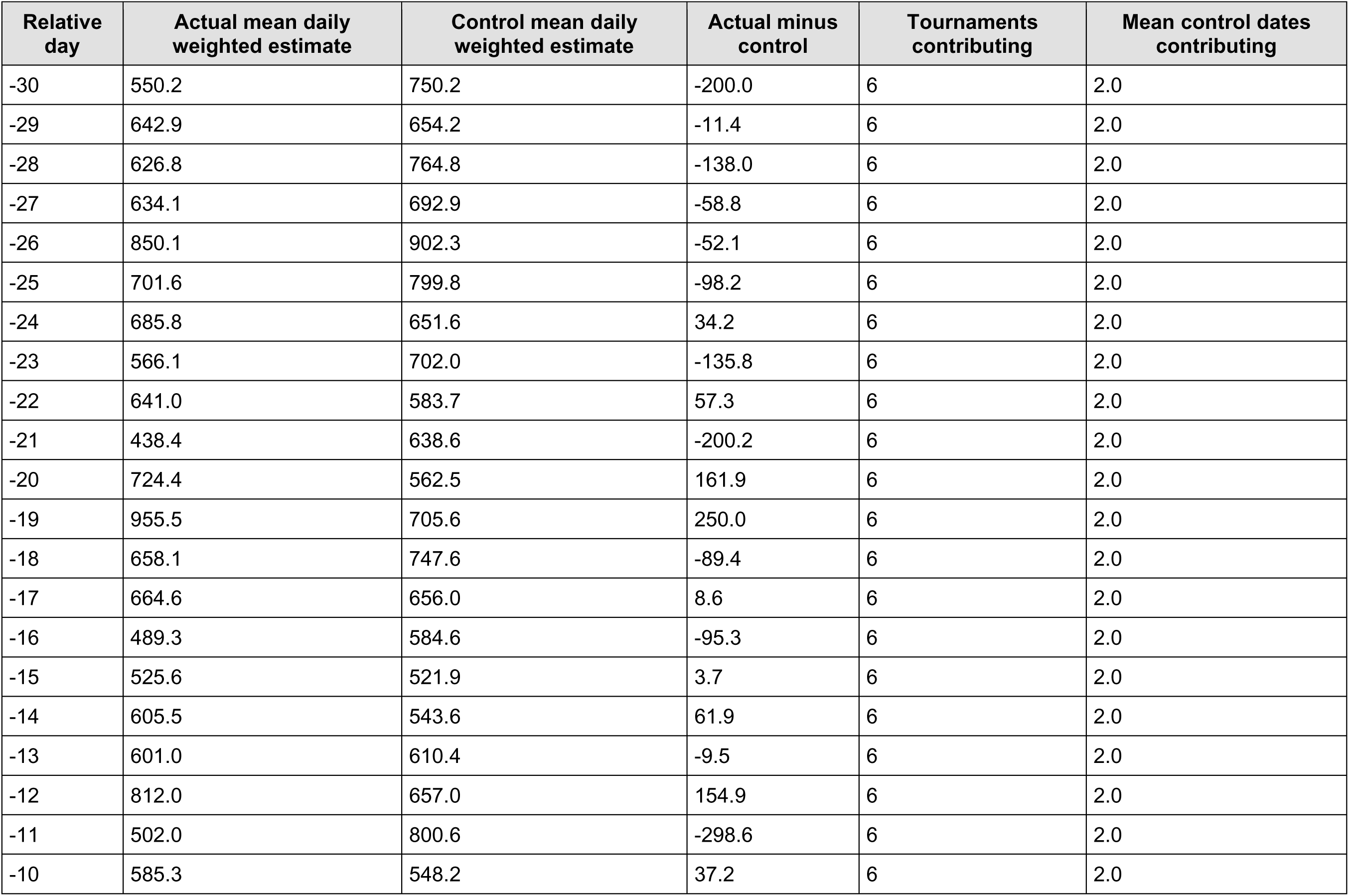
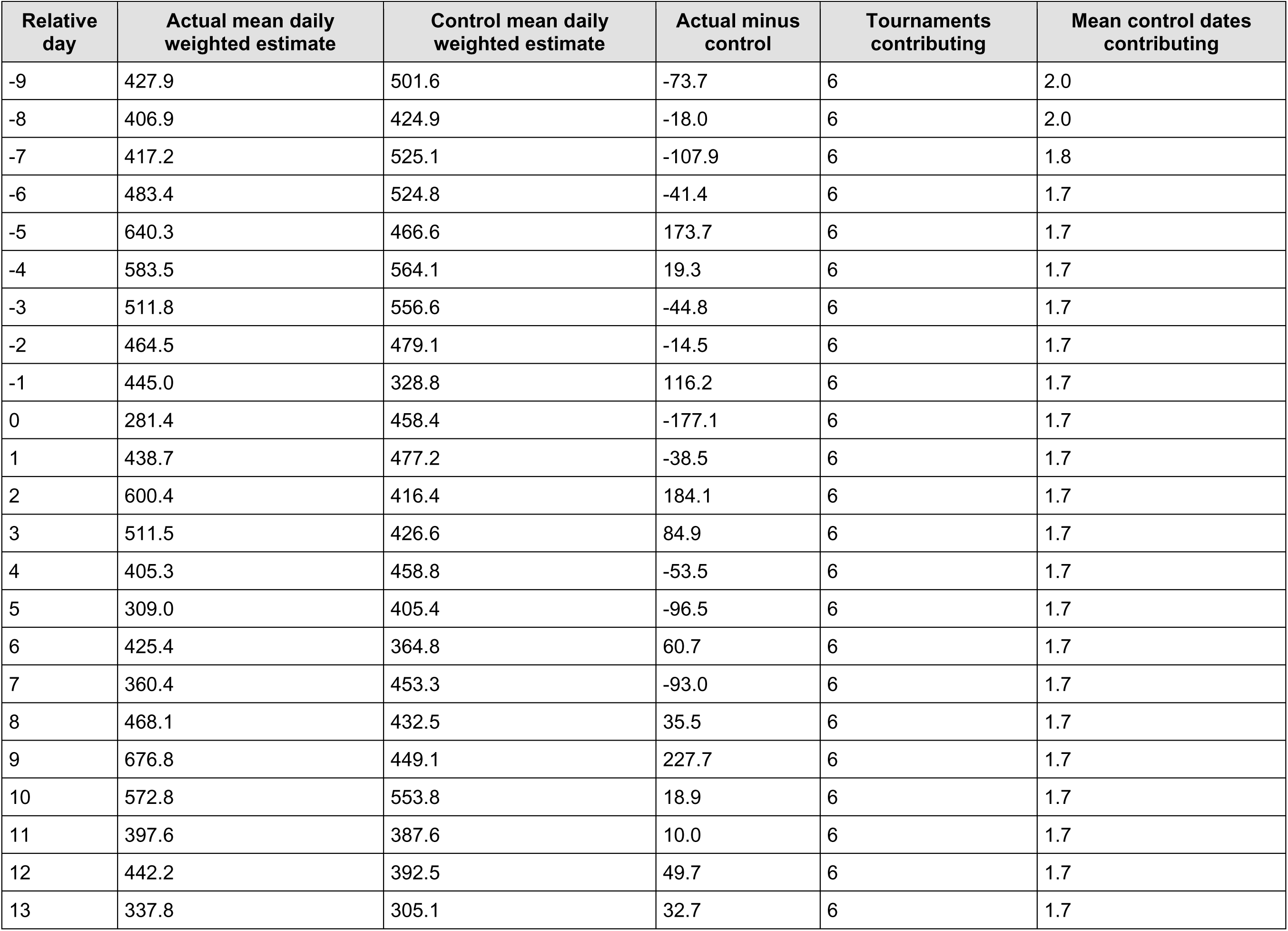
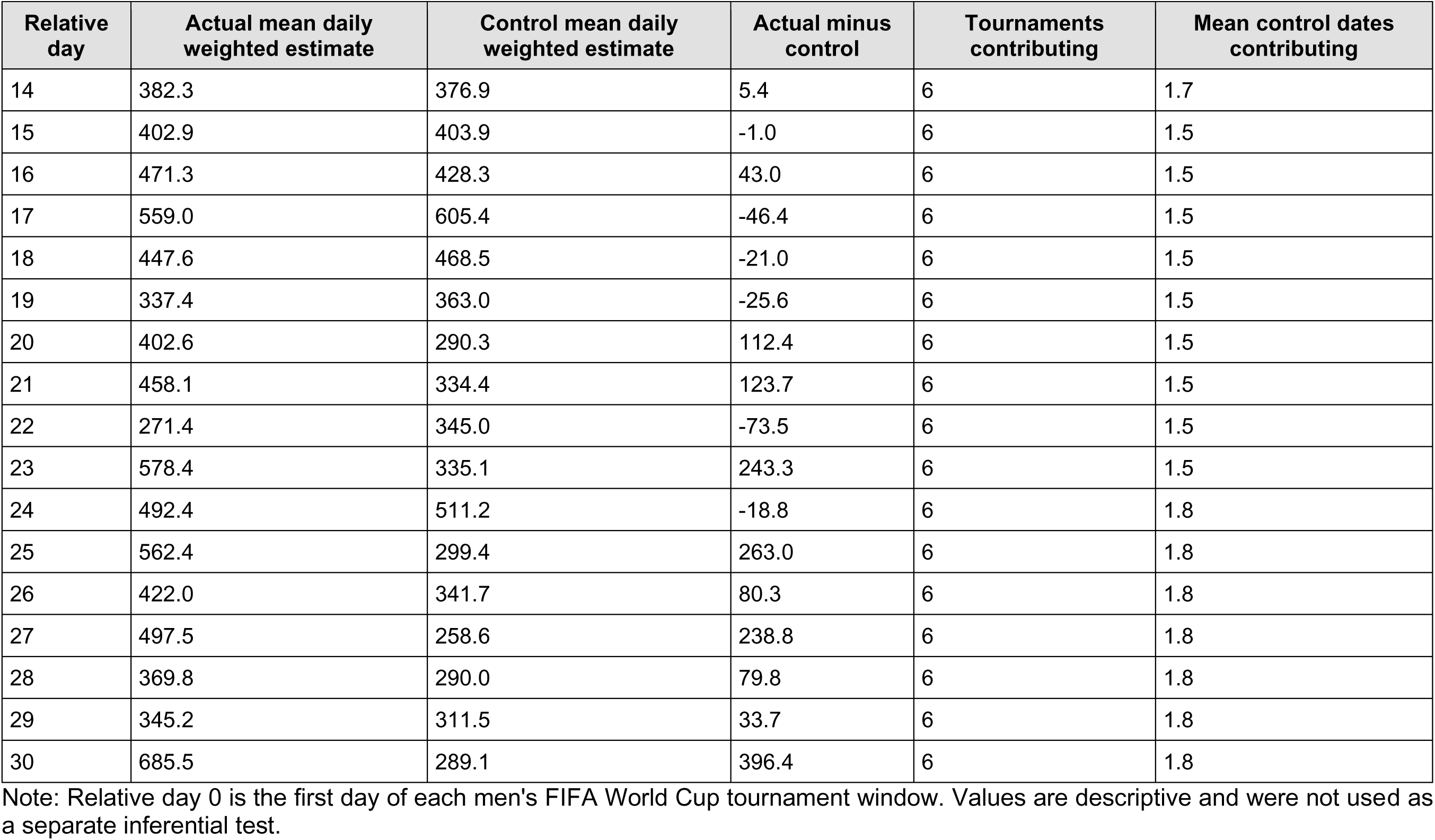
Daily values underlying the descriptive event-window profile.

| Relative day | Actual mean daily weighted estimate | Control mean daily weighted estimate | Actual minus control | Tournaments contributing | Mean control dates contributing |
| --- | --- | --- | --- | --- | --- |
| -30 | 550.2 | 750.2 | -200.0 | 6 | 2.0 |
| -29 | 642.9 | 654.2 | -11.4 | 6 | 2.0 |
| -28 | 626.8 | 764.8 | -138.0 | 6 | 2.0 |
| -27 | 634.1 | 692.9 | -58.8 | 6 | 2.0 |
| -26 | 850.1 | 902.3 | -52.1 | 6 | 2.0 |
| -25 | 701.6 | 799.8 | -98.2 | 6 | 2.0 |
| -24 | 685.8 | 651.6 | 34.2 | 6 | 2.0 |
| -23 | 566.1 | 702.0 | -135.8 | 6 | 2.0 |
| -22 | 641.0 | 583.7 | 57.3 | 6 | 2.0 |
| -21 | 438.4 | 638.6 | -200.2 | 6 | 2.0 |
| -20 | 724.4 | 562.5 | 161.9 | 6 | 2.0 |
| -19 | 955.5 | 705.6 | 250.0 | 6 | 2.0 |
| -18 | 658.1 | 747.6 | -89.4 | 6 | 2.0 |
| -17 | 664.6 | 656.0 | 8.6 | 6 | 2.0 |
| -16 | 489.3 | 584.6 | -95.3 | 6 | 2.0 |
| -15 | 525.6 | 521.9 | 3.7 | 6 | 2.0 |
| -14 | 605.5 | 543.6 | 61.9 | 6 | 2.0 |
| -13 | 601.0 | 610.4 | -9.5 | 6 | 2.0 |
| -12 | 812.0 | 657.0 | 154.9 | 6 | 2.0 |
| -11 | 502.0 | 800.6 | -298.6 | 6 | 2.0 |
| -10 | 585.3 | 548.2 | 37.2 | 6 | 2.0 |
| -9 | 427.9 | 501.6 | -73.7 | 6 | 2.0 |
| -8 | 406.9 | 424.9 | -18.0 | 6 | 2.0 |
| -7 | 417.2 | 525.1 | -107.9 | 6 | 1.8 |
| -6 | 483.4 | 524.8 | -41.4 | 6 | 1.7 |
| -5 | 640.3 | 466.6 | 173.7 | 6 | 1.7 |
| -4 | 583.5 | 564.1 | 19.3 | 6 | 1.7 |
| -3 | 511.8 | 556.6 | -44.8 | 6 | 1.7 |
| -2 | 464.5 | 479.1 | -14.5 | 6 | 1.7 |
| -1 | 445.0 | 328.8 | 116.2 | 6 | 1.7 |
| 0 | 281.4 | 458.4 | -177.1 | 6 | 1.7 |
| 1 | 438.7 | 477.2 | -38.5 | 6 | 1.7 |
| 2 | 600.4 | 416.4 | 184.1 | 6 | 1.7 |
| 3 | 511.5 | 426.6 | 84.9 | 6 | 1.7 |
| 4 | 405.3 | 458.8 | -53.5 | 6 | 1.7 |
| 5 | 309.0 | 405.4 | -96.5 | 6 | 1.7 |
| 6 | 425.4 | 364.8 | 60.7 | 6 | 1.7 |
| 7 | 360.4 | 453.3 | -93.0 | 6 | 1.7 |
| 8 | 468.1 | 432.5 | 35.5 | 6 | 1.7 |
| 9 | 676.8 | 449.1 | 227.7 | 6 | 1.7 |
| 10 | 572.8 | 553.8 | 18.9 | 6 | 1.7 |
| 11 | 397.6 | 387.6 | 10.0 | 6 | 1.7 |
| 12 | 442.2 | 392.5 | 49.7 | 6 | 1.7 |
| 13 | 337.8 | 305.1 | 32.7 | 6 | 1.7 |
| 14 | 382.3 | 376.9 | 5.4 | 6 | 1.7 |
| 15 | 402.9 | 403.9 | -1.0 | 6 | 1.5 |
| 16 | 471.3 | 428.3 | 43.0 | 6 | 1.5 |
| 17 | 559.0 | 605.4 | -46.4 | 6 | 1.5 |
| 18 | 447.6 | 468.5 | -21.0 | 6 | 1.5 |
| 19 | 337.4 | 363.0 | -25.6 | 6 | 1.5 |
| 20 | 402.6 | 290.3 | 112.4 | 6 | 1.5 |
| 21 | 458.1 | 334.4 | 123.7 | 6 | 1.5 |
| 22 | 271.4 | 345.0 | -73.5 | 6 | 1.5 |
| 23 | 578.4 | 335.1 | 243.3 | 6 | 1.5 |
| 24 | 492.4 | 511.2 | -18.8 | 6 | 1.8 |
| 25 | 562.4 | 299.4 | 263.0 | 6 | 1.8 |
| 26 | 422.0 | 341.7 | 80.3 | 6 | 1.8 |
| 27 | 497.5 | 258.6 | 238.8 | 6 | 1.8 |
| 28 | 369.8 | 290.0 | 79.8 | 6 | 1.8 |
| 29 | 345.2 | 311.5 | 33.7 | 6 | 1.8 |
| 30 | 685.5 | 289.1 | 396.4 | 6 | 1.8 |

Note: Relative day 0 is the first day of each men's FIFA World Cup tournament window. Values are descriptive and were not used as a separate inferential test.

**Supplementary Table S9.** Narrative-keyword and combined-definition audit by year.

| Year | Definition | Unweighted n | Weighted estimate | 95% CI | CV | Weighted % | Denominator weighted estimate |
| --- | --- | --- | --- | --- | --- | --- | --- |
| 1999 | soccer_narrative | 4,493 | 173,446 | 135,276 to 211,617 | 0.112 | 1.43% | 12,165,225 |
| 1999 | soccer_combined | 4,561 | 176,131 | 136,886 to 215,375 | 0.114 | 1.45% | 12,165,225 |
| 2000 | soccer_narrative | 4,797 | 183,020 | 134,934 to 231,106 | 0.134 | 1.42% | 12,924,488 |
| 2000 | soccer_combined | 4,873 | 185,532 | 136,641 to 234,423 | 0.134 | 1.44% | 12,924,488 |
| 2001 | soccer_narrative | 4,787 | 173,143 | 140,132 to 206,153 | 0.097 | 1.30% | 13,310,088 |
| 2001 | soccer_combined | 4,867 | 176,286 | 143,076 to 209,497 | 0.096 | 1.32% | 13,310,088 |
| 2002 | soccer_narrative | 4,859 | 168,274 | 135,047 to 201,502 | 0.101 | 1.32% | 12,790,786 |
| 2002 | soccer_combined | 4,983 | 173,986 | 139,222 to 208,750 | 0.102 | 1.36% | 12,790,786 |
| 2003 | soccer_narrative | 4,588 | 165,170 | 127,982 to 202,358 | 0.115 | 1.30% | 12,720,947 |
| 2003 | soccer_combined | 4,667 | 167,594 | 129,838 to 205,349 | 0.115 | 1.32% | 12,720,947 |
| 2004 | soccer_narrative | 4,866 | 172,225 | 131,293 to 213,156 | 0.121 | 1.31% | 13,096,983 |
| 2004 | soccer_combined | 4,933 | 174,168 | 132,884 to 215,453 | 0.121 | 1.33% | 13,096,983 |
| 2005 | soccer_narrative | 5,223 | 173,104 | 132,181 to 214,028 | 0.121 | 1.37% | 12,608,693 |
| 2005 | soccer_combined | 5,306 | 175,198 | 133,771 to 216,625 | 0.121 | 1.39% | 12,608,693 |
| 2006 | soccer_narrative | 5,481 | 185,313 | 141,227 to 229,399 | 0.121 | 1.40% | 13,232,263 |
| 2006 | soccer_combined | 5,532 | 187,079 | 142,632 to 231,526 | 0.121 | 1.41% | 13,232,263 |
| 2007 | soccer_narrative | 5,984 | 198,043 | 150,669 to 245,417 | 0.122 | 1.50% | 13,232,338 |
| 2007 | soccer_combined | 6,033 | 199,752 | 152,076 to 247,428 | 0.122 | 1.51% | 13,232,338 |
| 2008 | soccer_narrative | 6,161 | 198,720 | 151,112 to 246,328 | 0.122 | 1.48% | 13,456,353 |
| 2008 | soccer_combined | 6,213 | 200,293 | 152,368 to 248,217 | 0.122 | 1.49% | 13,456,353 |
| 2009 | soccer_narrative | 6,584 | 206,648 | 161,363 to 251,933 | 0.112 | 1.48% | 13,966,898 |
| 2009 | soccer_combined | 6,653 | 209,063 | 163,396 to 254,730 | 0.111 | 1.50% | 13,966,898 |
| 2010 | soccer_narrative | 7,203 | 223,599 | 175,380 to 271,818 | 0.110 | 1.52% | 14,694,928 |
| 2010 | soccer_combined | 7,289 | 226,864 | 178,553 to 275,175 | 0.109 | 1.54% | 14,694,928 |
| 2011 | soccer_narrative | 6,867 | 208,468 | 160,371 to 256,565 | 0.118 | 1.47% | 14,162,084 |
| 2011 | soccer_combined | 7,025 | 215,051 | 166,983 to 263,119 | 0.114 | 1.52% | 14,162,084 |
| 2012 | soccer_narrative | 7,275 | 227,801 | 173,865 to 281,737 | 0.121 | 1.56% | 14,614,128 |
| 2012 | soccer_combined | 7,377 | 232,507 | 178,083 to 286,931 | 0.119 | 1.59% | 14,614,128 |
| 2013 | soccer_narrative | 7,212 | 225,628 | 166,987 to 284,270 | 0.133 | 1.61% | 14,033,745 |
| 2013 | soccer_combined | 7,314 | 230,353 | 171,321 to 289,385 | 0.131 | 1.64% | 14,033,745 |
| 2014 | soccer_narrative | 7,532 | 238,361 | 169,791 to 306,931 | 0.147 | 1.72% | 13,860,971 |
| 2014 | soccer_combined | 7,609 | 240,897 | 172,022 to 309,772 | 0.146 | 1.74% | 13,860,971 |
| 2015 | soccer_narrative | 7,201 | 224,915 | 159,016 to 290,815 | 0.149 | 1.59% | 14,132,697 |
| 2015 | soccer_combined | 7,282 | 228,547 | 161,660 to 295,434 | 0.149 | 1.62% | 14,132,697 |
| 2016 | soccer_narrative | 7,619 | 223,784 | 151,805 to 295,763 | 0.164 | 1.56% | 14,319,418 |
| 2016 | soccer_combined | 7,708 | 227,326 | 153,925 to 300,727 | 0.165 | 1.59% | 14,319,418 |
| 2017 | soccer_narrative | 7,496 | 215,455 | 155,773 to 275,138 | 0.141 | 1.46% | 14,727,704 |
| 2017 | soccer_combined | 7,589 | 219,750 | 159,030 to 280,469 | 0.141 | 1.49% | 14,727,704 |
| 2018 | soccer_narrative | 6,766 | 197,761 | 141,944 to 253,578 | 0.144 | 1.42% | 13,959,174 |
| 2018 | soccer_combined | 6,826 | 199,938 | 143,449 to 256,427 | 0.144 | 1.43% | 13,959,174 |
| 2019 | soccer_narrative | 6,598 | 187,387 | 131,752 to 243,021 | 0.151 | 1.39% | 13,464,372 |
| 2019 | soccer_combined | 6,637 | 189,310 | 133,651 to 244,969 | 0.150 | 1.41% | 13,464,372 |
| 2020 | soccer_narrative | 3,186 | 80,898 | 52,160 to 109,636 | 0.181 | 0.74% | 10,994,077 |
| 2020 | soccer_combined | 3,213 | 82,161 | 53,421 to 110,901 | 0.178 | 0.75% | 10,994,077 |
| 2021 | soccer_narrative | 5,620 | 144,388 | 104,889 to 183,887 | 0.140 | 1.23% | 11,738,091 |
| 2021 | soccer_combined | 5,646 | 145,669 | 106,171 to 185,167 | 0.138 | 1.24% | 11,738,091 |
| 2022 | soccer_narrative | 6,303 | 178,229 | 125,049 to 231,409 | 0.152 | 1.41% | 12,663,028 |
| 2022 | soccer_combined | 6,343 | 180,309 | 127,085 to 233,533 | 0.151 | 1.42% | 12,663,028 |
| 2023 | soccer_narrative | 7,577 | 211,123 | 152,828 to 269,417 | 0.141 | 1.66% | 12,740,256 |
| 2023 | soccer_combined | 7,639 | 214,111 | 155,539 to 272,682 | 0.140 | 1.68% | 12,740,256 |
| 2024 | soccer_narrative | 8,370 | 265,415 | 207,777 to 323,054 | 0.111 | 1.76% | 15,068,979 |
| 2024 | soccer_combined | 8,399 | 267,410 | 209,038 to 325,781 | 0.111 | 1.77% | 15,068,979 |
| 2025 | soccer_narrative | 8,998 | 265,811 | 200,937 to 330,684 | 0.125 | 1.61% | 16,485,413 |
| 2025 | soccer_combined | 9,003 | 265,975 | 201,130 to 330,820 | 0.124 | 1.61% | 16,485,413 |

Not: Narrative-derived definitions were used only as exploratory sensitivity outcomes.

**Supplementary Figure S1.**
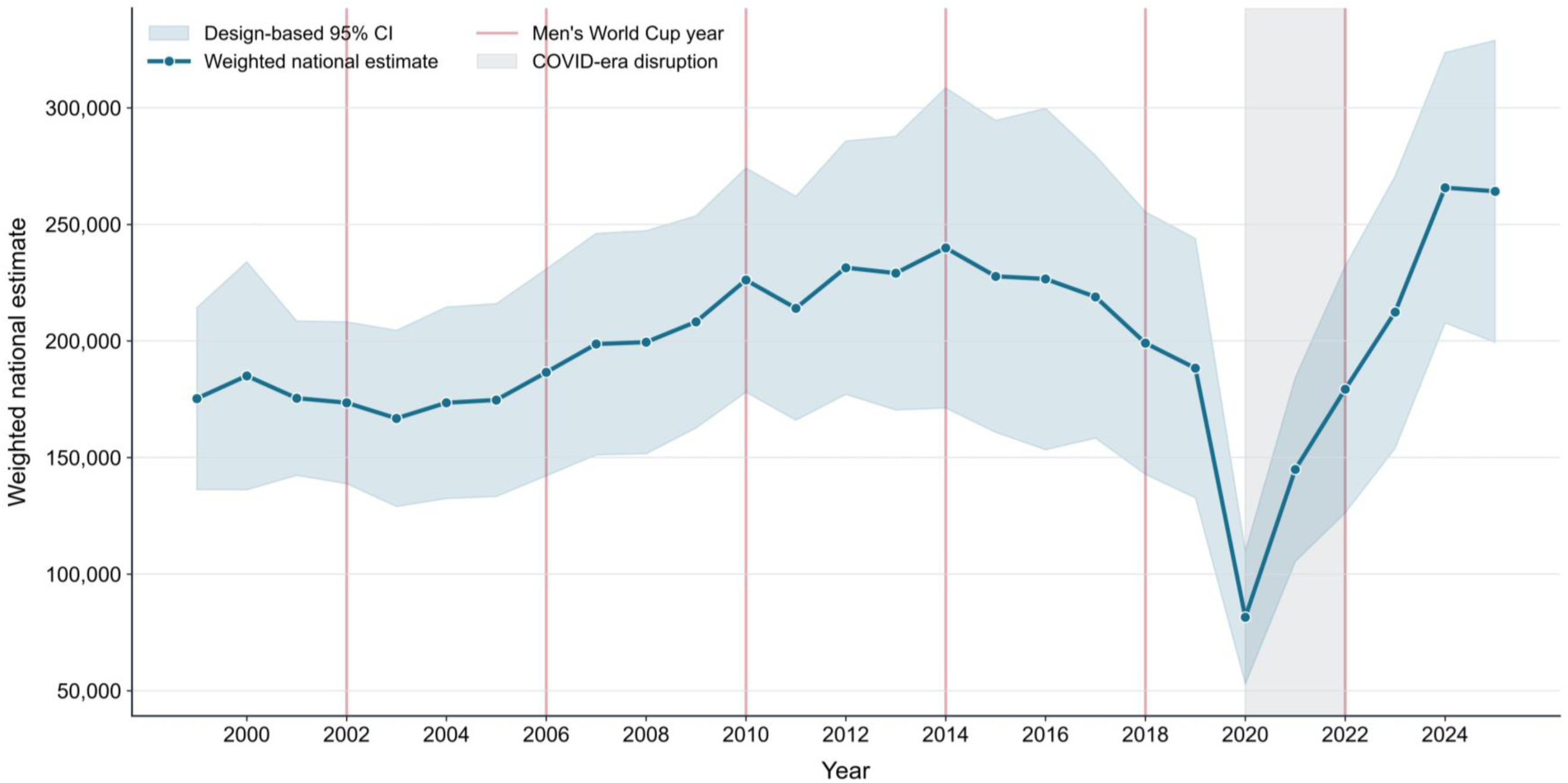
Annual weighted national estimates of ED-treated soccer-coded injuries with design-based 95% confidence intervals, men’s World Cup years marked, and COVID-era disruption indicated, NEISS 1999-2025.

**Supplementary Figure S2.**
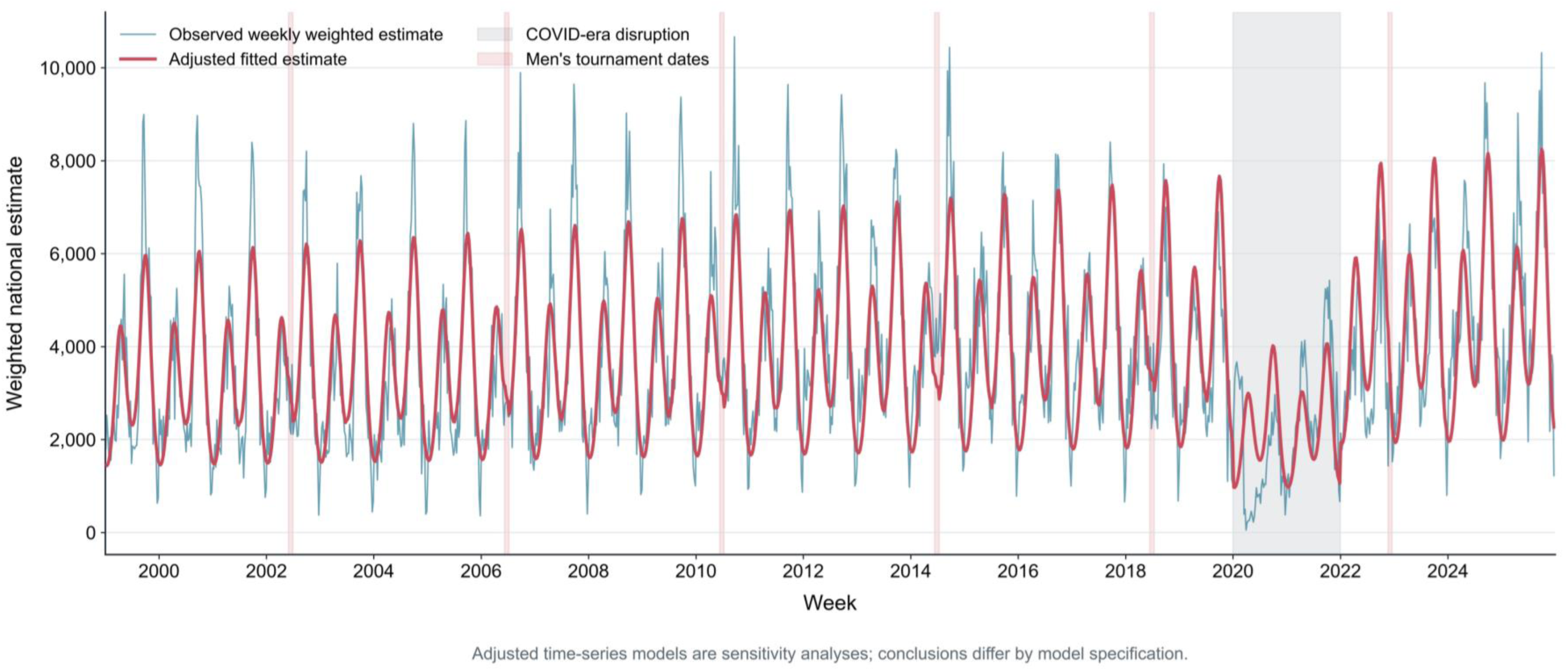
Weekly observed and fitted product-code-defined soccer injury estimates from the adjusted time-series sensitivity analysis. Adjusted models are presented as sensitivity analyses because residual dependence persisted and interval conclusions differed by specification.

**Supplementary Figure S3.**
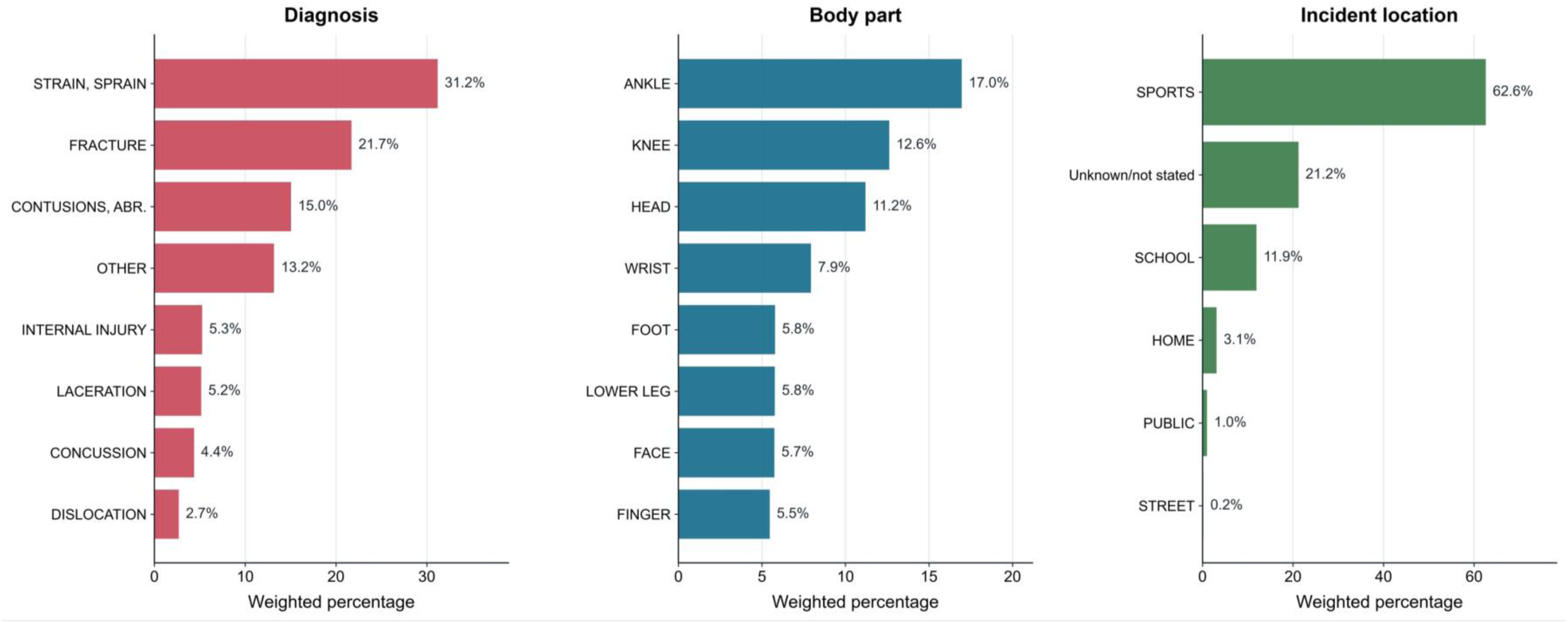
Weighted percentage distribution of stable diagnosis, body-part, and incident-location categories within the product-code-defined soccer cohort, NEISS 1999-2025. Bars are limited to categories meeting CPSC stability criteria; unstable categories are listed with suppressed weighted estimates in Table 2.

